# Co-creating the Butterfly Multimedia Patient Education Platform for Thyroid Surgery Along the Patient Pathway: Lessons for Participatory Digital Patient Education

**DOI:** 10.64898/2026.06.25.26356545

**Authors:** Maria Moschofidou, Gerasimos P. Sykiotis

**Affiliations:** Lausanne University Hospital and University of Lausanne, Lausanne Switzerland

**Keywords:** Digital patient education, participatory design, multimedia learning, thyroid surgery, shared decision-making, health literacy, co-creation

## Abstract

**Objective:** To describe and critically appraise the participatory design process underpinning a multimedia patient education platform for thyroid surgery in light of subsequent evaluation findings, in order to derive humanl71factors and methodological lessons for future col71creation.

**Methods:** Using a multi-phase participatory design approach anchored in the Technology Acceptance Model and Mayer’s Cognitive Theory of Multimedia Learning, including duall71channel processing and cognitive load principles, a multidisciplinary team including a patient representative col71created Medtronic’s Butterfly platform for preoperative thyroid patients. Two Nominal Group Technique-based workshops mapped a six-stage surgical pathway, identified stage-specific information needs, and specified formats and sequencing for educational content. An external communication agency developed storyboards as precursors to webpages, animated videos, and leaflets, which underwent iterative advisory board review and content validity indexing before external mixed-methods evaluation in a cohort of thyroidectomy candidates.

**Results:** The co-creation process yielded a modular platform comprising five informational webpages, five short animated videos, and three downloadable leaflets, each mapped to specific pathway stages, learning objectives, and information needs. The platform achieved high scores on validated measures of information quality, credibility, understandability, and actionability, and patients reported enhanced preparedness and decision-making support. However, formal readability assessments of webpages and video scripts showed that most materials exceeded recommended grade levels, usability issues in digital user experience (e.g., mobile responsiveness, navigation, subtitles) emerged only after implementation, and group-level stress, anxiety, and depression scores did not substantially improve despite perceived informational and emotional support.

**Conclusions:** Participatory design facilitated the creation of a clinically credible, pathway-aligned educational platform that addresses key informational gaps in thyroid surgery care but also exposed boundaries of information-focused co-creation. Future projects should treat readability as a non-negotiable design constraint, extend co-creation to systematic usability testing of digital interfaces, and explicitly distinguish educational from psychosocial aims while co-designing complementary distress screening and referral pathways. The Butterfly platform offers a transferable model for co-creating high-quality digital patient education while highlighting the need to more fully center equity, accessibility, and broader patient support in participatory design.

## INTRODUCTION

Access to comprehensive, accurate, and up-to-date information is essential to support informed patient decision-making and is crucial for enhancing clinical outcomes, improving patient experience, and increasing treatment adherence [1–3]. Notably, a systematic review of 176 studies demonstrated that patient education enhances pre-treatment decision-making, side-effect management, and treatment adherence, suggesting that effective patient education extends beyond reducing anxiety to actively supporting self-management throughout the care continuum [4]. In recent years, patients increasingly seek online health information, profoundly influencing their decisions and behaviors; nevertheless, many still lack adequate understanding of their available treatment options [5]. This gap is particularly evident in thyroid-related health information, where systematic reviews have identified that many websites provide outdated information misaligned with internationally endorsed guidelines, omit important treatment alternatives, and/or are written at readability levels exceeding average patients’ comprehension [6–8]. These accessibility barriers can have direct negative consequences for patients requiring thyroid treatment, such as surgical intervention. Indeed, for patients contemplating thyroid surgery, information-related stakes are particularly high, and web-based preoperative patient education has been shown to significantly reduce surgery-related anxiety in such patients and to enhance early postoperative recovery outcomes [9].

Video-based education appears particularly effective in addressing information gaps, as it outperforms traditional methods for patient learning by facilitating the visualization of complex processes and procedural preparation [10, 11]. Animated videos can simplify complex medical concepts, be adapted to different literacy levels, and enhance long-term information retention [4, 12, 13]. A meta-analysis of 21 studies reported that animated videos substantially improve patient learning outcomes compared with standard educational approaches, with particularly strong effects in clinical settings such as surgery [14]. These improvements appear driven by design principles grounded in cognitive learning theory: animations that reduce extraneous cognitive load through strategic visual cuing, clear narration, minimal background detail, and selective movement facilitate deeper processing and retention [15, 16]. Therefore, animated videos represent a promising format for addressing the literacy barriers that currently limit patient understanding of thyroid surgery options and related care.

Regardless of format, the mere availability of high-quality educational content does not guarantee its relevance or uptake. To be effective, educational materials must not only address appropriate literacy levels, provide clear and evidence-based content, and incorporate multimedia elements for complex concepts, but also, crucially, involve patients together with other end-users and stakeholders in the co-creation and evaluation of content [17, 18]. The latter ensures that real informational needs and preferences are addressed, supporting active patient participation and self-management. Unlike traditional top-down development, which relies on expert assumptions that may not match patients’ real priorities and communication preferences, participatory design actively involves patients and other stakeholders in creating health interventions so that materials are truly relevant and usable [17, 19, 20]. Clear guidance on how to co-create effectively is crucial for developing patient-centered interventions, and research has identified evidence-based best practices for co-creation; these include grounding work in existing literature, using an explicit theoretical framework, involving patients and clinicians from the start of development, ensuring diverse representation across stakeholder groups, distributing decision-making authority equitably among partners, and applying validated evaluation tools [17].

Building on these evidence-based principles for effective patient education and meaningful co-creation [17], we previously developed a comprehensive audiovisual information platform – Medtronic’s Butterfly platform – designed to support shared decision-making for patients with surgical thyroid disease (link available from Medtronic upon request). This new patient information material was purposefully designed by incorporating the key concepts and principles identified in the literature: it addresses varying literacy levels through multimodal content, provides evidence-based information aligned with current clinical guidelines [21, 22], utilizes animated video to enhance comprehension and retention, and was iteratively refined through clinician and patient feedback and involvement, including formal assessment in a mixed-methods study that demonstrated its perceived usefulness and impact. As a follow-up to the Butterfly platform’s recently published external validation study [23], the present manuscript documents the co-creation process that underpinned its development and offers a detailed model that can inform the design of similar patient-centered educational materials across disease areas. By elucidating how participatory design principles translate into practice – from stakeholder engagement and iterative prototyping through validation with real-life patient cohorts – this work aims to provide an actionable template for developing high-quality, patient-centered health information resources that genuinely reflect patient needs and preferences.

## METHODS

### Theoretical framework

The education platform for preoperative thyroid patients was developed through a structured, multi-phase process that combined participatory design principles with evidence-based content creation methods. Participatory design engages end-users and stakeholders collaboratively to co-create innovative products, such as data visualizations or health tools, ensuring relevance and usability through their direct involvement [24, 25]. The theoretical framework drew upon two complementary models. First, the technology acceptance model [16, 26] guided design decisions by focusing on maximizing patients’ perceived usefulness – defined as clear clinical benefits, anxiety reduction, and support for decision-making – and perceived ease of understanding, achieved through plain language, concise videos, and an intuitive structure. The multimedia components were designed in line with Mayer’s Cognitive Theory of Multimedia Learning, which posits that learners process information through dual verbal and visual channels with limited capacity and engage in active cognitive processing to integrate new information with prior knowledge. To translate these principles into practice, the platform distributed information across text, narration, and imagery and sought to minimize extraneous cognitive load through clear signaling, streamlined visuals, and avoidance of unnecessary detail, an approach consistent with recommendations for educational video design in clinical teaching [27, 28].

### Stakeholder representation and role distribution

Key stakeholder groups were purposefully selected to ensure comprehensive representation across clinical, patient, academic, and technological domains. Participants internal to Medtronic included a head and neck therapy manager who served as the project lead, a senior business director for ear, nose and throat (ENT), a director of reimbursement and health economics, a consultant specializing in optimizing hospital care pathways and health service delivery, a senior market developer specialist for head and neck surgery, a product manager for ENT, and a health communication specialist writer. Together, these internal stakeholders contributed expertise in clinical therapy areas, health solutions and educational content design, market access, and strategic implementation.

External participants, forming an advisory board, included three endocrine surgeons, two endocrinologists, and one patient representative from a national patient association. Additionally, commercial digital health experts were commissioned to prepare the videos and accompanying text of the audiovisual material, and a doctoral student was assigned to perform the external validation study with real-life patients. This multidisciplinary composition ensured that the development process integrated clinical expertise with user-centered and evidence-based perspectives, aiming for a relevant, credible, and actionable educational resource.

Specific contributors from each stakeholder category were appointed based on their expertise and function (**Figure 1**). The medical content was co-designed and validated by surgeons and endocrinologists in collaboration with the project lead and other Medtronic team members, with experiential insights and communication preferences provided by the patient representative. Linguistic clarity, narrative coherence, and targeted review of supporting literature were ensured by the health communication specialist. Digital health experts created and optimized the platform’s architecture, accessibility, and interactivity. The doctoral student conducted the cohort study supporting external validation of the developed educational resources, under supervision by one of the participating endocrinologists. The Medtronic project leader coordinated interprofessional teams, managed project timelines, and organized structural tasks, including the facilitation of the co-creation workshops and between-workshop participant feedback loops. During the co-creation process, stakeholders collaborated iteratively to map patient experiences and define critical educational priorities, with feedback loops guiding content refinement to ensure clinical accuracy, clarity, and usability.

**Figure 1.**
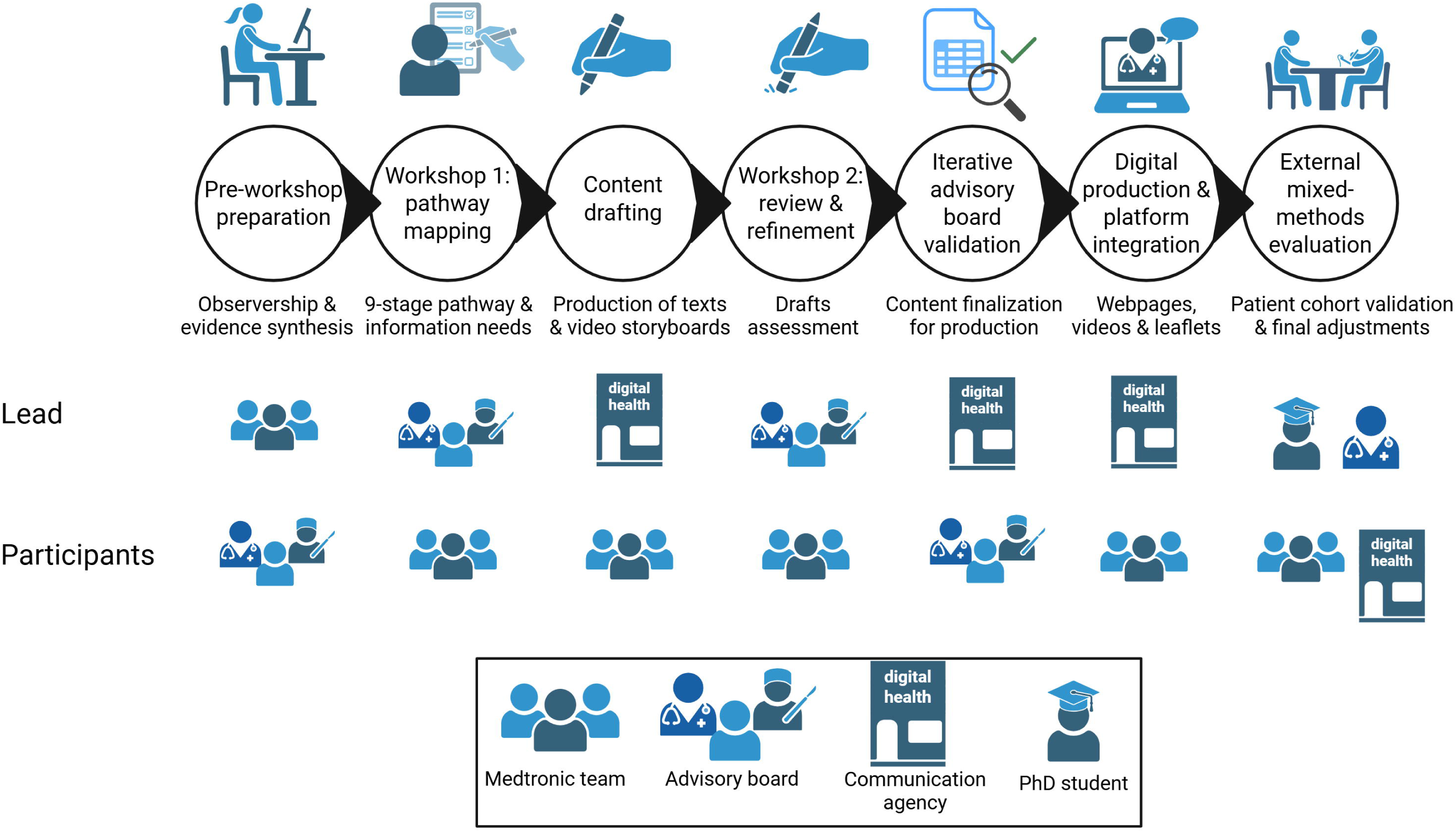
Cocreation process and stakeholder involvement. Development phases and stakeholder roles (lead and participant) in creating the Butterfly multimedia patient education platform for thyroid surgery.

### Pre-workshop formative activities

To prepare for the project, pre-workshop formative activities were undertaken to ground the co-creation process in both real-world clinical practice and current evidence. The Medtronic project manager completed an observership of four presurgical patient consultations with a high-volume thyroid surgeon in a tertiary academic medical center, to understand typical information flows, patient questions, and decision points in routine care. In parallel, the Medtronic medical writer conducted a targeted literature review on patient education best practices and thyroid surgery information needs and synthesized key findings into a summary that was shared with all participants before the first workshop. Together, these preparatory activities provided a shared clinical and evidence-based context for the stakeholder discussions and helped ensure that subsequent co-creation work was anchored in both observed practice and existing literature, consistent with recommendations for integrating health literacy and participatory design in patient-facing materials [17, 18]. The participating clinicians were all thyroid experts, well-versed in international guidelines in the field.

### Outline of the co-creation process

The co-creation process centered on two facilitated in-person workshops held eight months apart, bringing together Medtronic stakeholders and the advisory board. To generate, discuss, and rank ideas collaboratively, the workshops used the Nominal Group Technique (NGT), a structured consensus method frequently applied in healthcare [29]. In these sessions, participants worked through cycles of independent idea generation, round-robin sharing without criticism, group clarification in alternating break-out and plenary modes, and collective voting. Medtronic representatives acted as both participants and facilitators throughout.

During the first workshop, the patient pathway was mapped, and experts identified critical information needs at each stage using post-it exercises, prioritization matrices, brainstorming, and presentations of typical patient journeys. This process defined the platform’s key content stages, formats, and timing, ensuring that materials addressed the most relevant patient concerns. The outcomes guided the structure and initial content outline for the new educational material.

Between the workshops, content was developed by an external communication agency (Outlook Creative, Northampton, UK; https://outlookcreative.uk/) specializing in patient-centered health communication. They translated workshop results into texts and video storyboards that combined concrete examples with experiential advice, converting key recommendations into practical guidance for patients. The second workshop focused on reviewing and finalizing the professional drafts, particularly the storyboards that preceded video production. Advisory board members completed online surveys beforehand to prioritize topics, highlight missing issues, and comment on draft content.

Following the workshops, the advisory board conducted iterative online reviews to further refine clarity and relevance until full consensus was reached. Each item was evaluated using a content validity index; those falling below predefined thresholds were revised. Following approval of the final storyboards, Outlook Creative produced the audiovisual materials. Advisory board members then reviewed the webpages, videos, and downloadable leaflets in a final remote improvement round. An external validation study, reported separately [23], later confirmed content quality and informed the last adjustments, including addition of a subsection on nutrition that was specifically requested by some patients (Section 4.4, **Table 1**).

### Readability and linguistic analyses of video transcripts

To complement the previously reported analyses of the webpages, the same procedures were applied to video transcripts. Readability was assessed using standard formulas, including Flesch Reading Ease (FRE) and grade-level indices, and linguistic features were analyzed using the Linguistic Inquiry and Word Count (LIWC-22) tool. All analyses followed the same methodology and reporting approach described previously [23]. Because the downloadable leaflets were primarily formatted as bullet points and fragmented text, they were not suitable for reliable readability and LIWC-22 linguistic analysis, which require continuous prose.

## RESULTS

### Patient pathway mapping and needs assessment

Building on the two co-creation workshops described in the Methods, the multidisciplinary team outlined a six-stage patient pathway to guide the development of all educational materials. The pathway encompassed key stages in the care of surgical thyroid disease, ranging from (i) initial detection and (ii) diagnostic workup, through (iii) surgical decision-making, (iv) hospital admission, surgery, and the immediate postoperative period, (v) postoperative care at home, and (vi) long-term follow-up, including any additional treatment (**Figure 2**).

**Figure 2.**
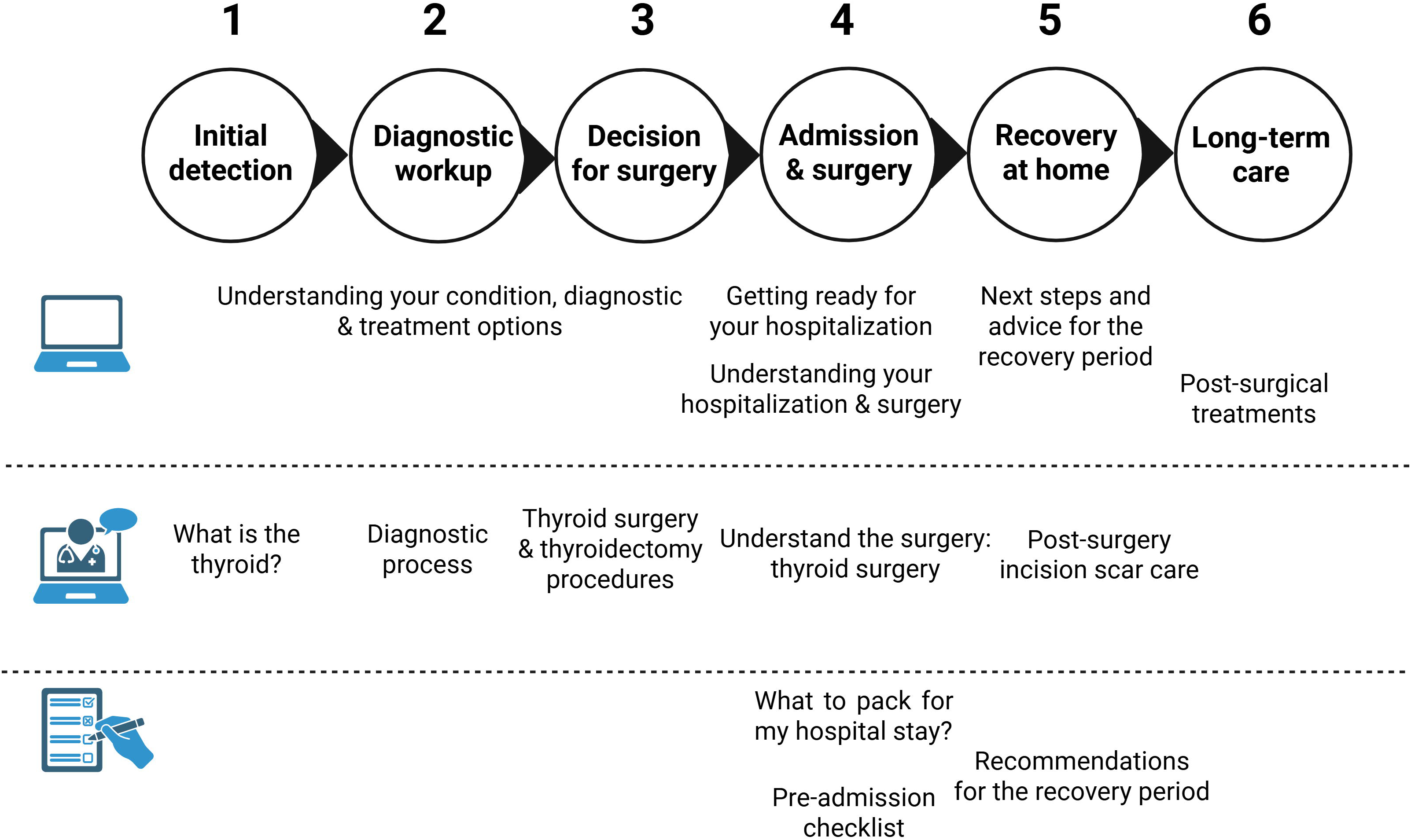
Patient pathway and mapping of webpages, videos, and leaflets. Alignment of Butterfly platform webpages, animated videos, and downloadable leaflets with the six-stage patient pathway for surgical thyroid disease.

For each stage, the team characterized the main healthcare actors, key examinations, potential support sources, and specific patient information needs, using triangulated input from surgeons, endocrinologists, a patient representative, and industry stakeholders. The mapping highlighted pronounced information gaps, particularly during the “unawareness to suspicion” phase, where patients seeking information online may encounter misleading or unreliable content, and the “diagnosis confirmation” stage, often marked by prolonged uncertainty and anxiety across multiple appointments.

This pathway also provided the conceptual foundation for structuring educational content, ensuring that information was available from the outset while organized into sequential, journey-aligned modules reflecting patient needs and key clinical decision/action points. The Butterfly platform was designed with a modular architecture comprising three complementary content formats: (i) five informational webpages organized into 18 subsections, (ii) five short animated videos, and (iii) three downloadable leaflets (**Figure 3**). **Tables 1-3** detail how each component addresses specific learning objectives and information needs mapped to corresponding stages of the patient journey, thereby operationalizing the participatory design consensus.

**Figure 3.**
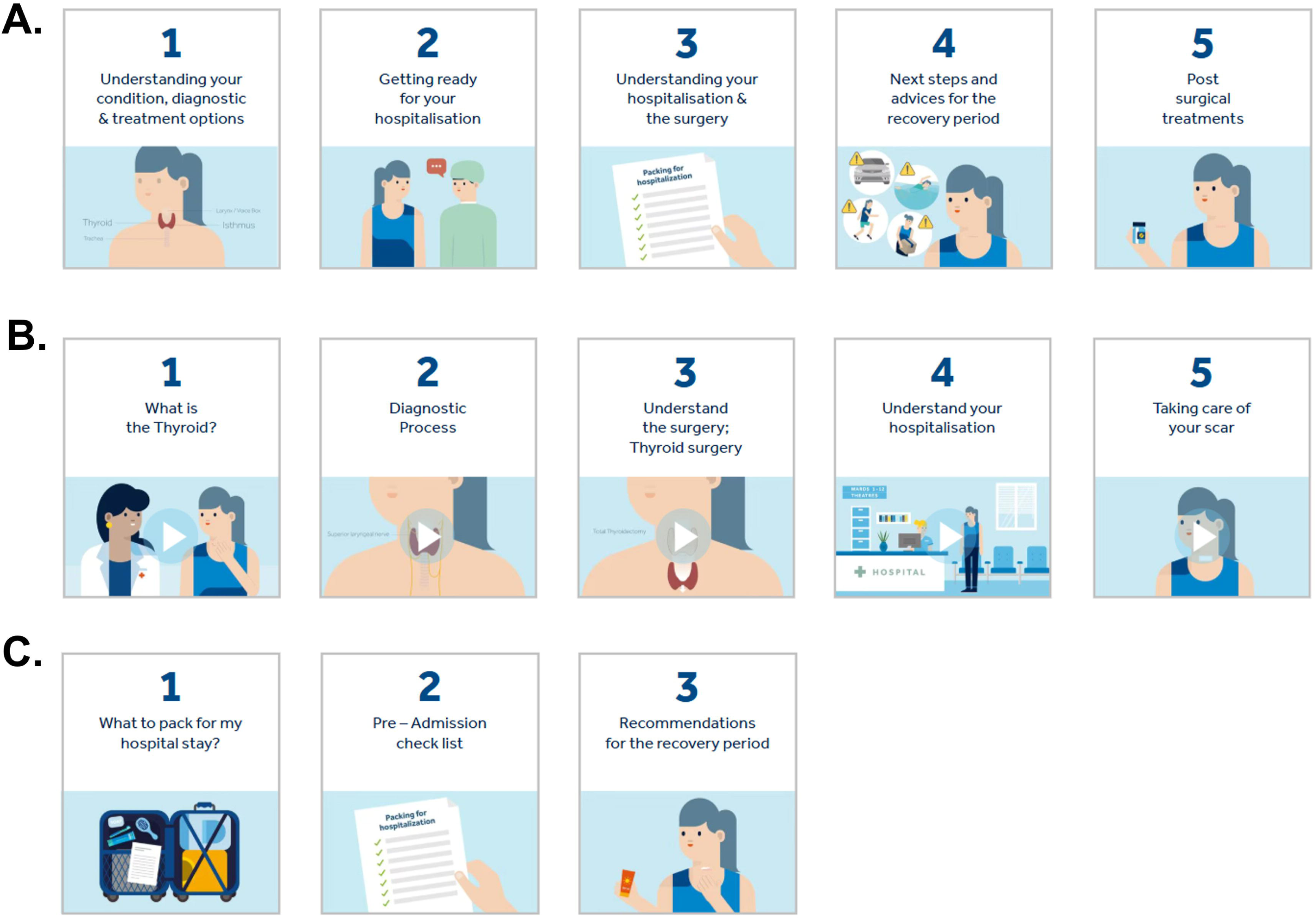
Screenshots of the Butterfly multimedia patient education platform main menus. **A.** Webpages. **B.** Videos. **C.** Downloadable leaflets.

### Informational webpages

Five main webpages were structured to mirror the patient pathway, from initial disease awareness through long-term follow-up. Each page contains multiple subsections (18 in total), providing evidence-based information in clear, patient-friendly language supported by visual aids. **Table 1** maps each subsection to its learning objectives, primary information needs addressed, corresponding stages of the patient journey, and, where relevant, associated videos and downloadable leaflets.

The content was initially adapted from pre-existing Medtronic patient education materials of more limited scope and subsequently expanded and refined through workshop feedback to address surgical risks, preoperative protocols, hospitalization experiences, postoperative care, and psychosocial support strategies.

### Animated educational videos

Five short animated videos (total duration range: 2:19 to 6:03 minutes; mean: 4:31) were developed and aligned with specific stages of the patient pathway. Video duration was defined based on total audiovisual length (including narration and background audio), consistent with evidence-based recommendations for patient education. Short, focused videos were chosen to align with evidence that learner engagement with educational videos drops markedly as length increases, with median engagement times highest for videos under about 6 minutes and substantially lower for longer formats [30]. This design is also consistent with cognitive load theory, which emphasizes segmenting information and limiting the amount of content processed at one time [31]. As shown in **Table 2**, each video targets a clearly defined information need with a limited set of learning objectives (maximum five per video), consistent with cognitive load theory [32] and the dual-channel design principles described in the Methods. The videos were designed to minimize extraneous cognitive load through strategic visual cueing, clear narration, and limited background detail, thereby facilitating comprehension and retention.

The consensus reading level of the video transcripts corresponded to approximately 9^th^-11th U.S. grade, exceeding recommended thresholds for patient-facing materials (**Supplementary Table 1**). FRE scores classified the texts as “fairly difficult” to “slightly difficult,” with videos 1-3 showing the highest linguistic complexity, and only modest reductions in videos 4 and 5. LIWC-22 analysis (**Supplementary Table 2**) confirmed good dictionary coverage and revealed clear stylistic differences across videos. The first three videos were more analytic and technically oriented, with more complex vocabulary and information-dense language, whereas the preparation and aftercare videos used simpler wording and a less formal structure. These latter videos also adopted a more interpersonal, patient-oriented style, with more direct address and greater use of social and supportive language, as well as a more reassuring tone. Despite this shift in discourse style, readability levels remained consistently high across all videos.

### Downloadable patient leaflets

Three one-page downloadable leaflets provide practical checklists and actionable guidance for hospital preparation and early recovery, corresponding to stages 1-4 of the patient pathway (**Table 3**). These leaflets operationalize the participatory design principle of providing “get-ready” resources that reduce cognitive burden by breaking complex clinical instructions into discrete, manageable tasks. Each leaflet addresses both clinical requirements and psychosocial aspects of care transitions, supporting patient agency and self-management.

### Integration and design coherence

The modular architecture of the Butterfly platform ensures that webpages, videos, and leaflets function as complementary rather than redundant resources. Webpages provide comprehensive, reference-style information accessible across the care continuum; videos deliver focused, easily digestible content targeting specific learning objectives and grounded in cognitive theory; and leaflets offer portable, just-in-time checklists for key transitions. Cross-referencing between formats (e.g., from webpage subsections to related videos) creates an integrated learning experience (**Table 1**, **Figure 2**). Τhis structural coherence reflects the participatory design process, in which stakeholders jointly defined not only what information to provide, but also how it should be delivered across the patient journey.

## DISCUSSION

### Overview and main contributions

This manuscript documents the participatory design process underpinning the development of the Butterfly platform, a comprehensive audiovisual educational resource for patients undergoing thyroid surgery. The co-creation workflow – grounded in the technology acceptance model and Mayer’s Cognitive Theory of Multimedia Learning, including dual-channel processing and cognitive load principles, operationalized through two structured workshops using the Nominal Group Technique (NGT), and refined through iterative multidisciplinary review – yielded a modular platform comprising five webpages, five animated videos, and three downloadable leaflets, systematically mapped to six stages of the surgical care continuum. External validation in a mixed-methods cohort study demonstrated that this process achieved core design objectives related to content quality, clinical credibility, structural coherence, and actionability, while also identifying areas for improvement in readability, digital user experience, and psychosocial impact [23].

### Content quality and clinical credibility through co-creation

The iterative pathway-mapping workshops and subsequent advisory board validation process directly translated stakeholder input into clinically accurate, decision-relevant content. Except for readability, formal evaluation using validated instruments covering usefulness, impact, understandability, and actionability confirmed that the platform exceeded predefined thresholds in all assessed domains [23]. Patient ratings of perceived usefulness further validated this approach, indicating that the co-creation workflow (NGT-based prioritization, expert content development by a health communication specialist, and multi-stakeholder review) effectively bridged the gap between clinical expertise and patient information needs. This outcome contrasts sharply with the documented deficiencies of many thyroid-related websites, which provide fragmented, outdated, or guideline-discordant information [6–8]. By embedding clinical and patient representatives throughout the design process rather than consulting them only retrospectively, the participatory approach ensured that alignment with current evidence and clinical workflows was built into the platform’s foundation rather than verified merely post hoc.

### From patient pathway mapping to actionable architecture

The six-stage patient pathway co-created during the first workshop provided the conceptual scaffold for the platform’s architecture, ensuring that content was structured along the patient journey and aligned with key clinical decision points and evolving patient needs. This pathway-driven structure distinguishes the Butterfly platform from many existing educational resources that typically present thyroid-related information in disease-centric or procedure-centric formats without explicit consideration of when patients encounter specific questions or anxieties [6, 33]. The resulting modular design of the platform, organized around “what–why–what next” principles, achieved high scores for understandability and actionability in the validation study, supporting patients in integrating information into preparation, decision-making, and self-management [23].

The patient pathway was conceptualized as a six-stage continuum from initial unawareness to long-term follow-up after thyroid surgery, beginning with incidental or nonspecific findings that trigger suspicion and diagnostic work-up, progressing through the “shock of diagnosis” and subsequent treatment decision-making, and continuing into perioperative care, early recovery at home, and long-term survivorship. Across these stages, patients experience shifting informational and psychosocial needs, including anxiety during diagnostic uncertainty, decisional complexity when weighing treatment options, stress around hospitalization and potential complications, and challenges related to recovery, hormone replacement, and “life without a thyroid” and/or “life after thyroid cancer.” This stage-based understanding informed the modular, journey-aligned architecture of the Butterfly platform, ensuring that educational content corresponded to the evolving questions, decisions, and self-management tasks patients face over time.

### Designing multimodal resources along the patient journey

Within this pathway-oriented architecture, the three content formats (webpages, videos, and downloadable leaflets) played distinct but complementary roles. The webpages provided comprehensive, reference-style information that patients could consult at any point, supporting in-depth understanding of disease mechanisms, diagnostic procedures, and treatment options. The animated videos translated complex concepts, such as surgical anatomy and perioperative workflows, into concise and intuitive visual narratives designed to enhance comprehension and recall. The downloadable leaflets functioned as transition tools, offering practical checklists and “get-ready” guidance around hospitalization and early recovery. Together, these multimodal resources operationalized the pathway mapping into an actionable educational ecosystem, with content elements mapped to specific stages yet fully accessible across the continuum of care.

### Designing patient education videos with dual-channel principles

The explicit application of Mayer’s Cognitive Theory of Multimedia Learning, including dual-channel processing and cognitive load principles, during video design represents a successful translation of cognitive learning principles into practice through participatory methods [15, 16, 32]. By distributing information across synchronized narration and animation, limiting learning objectives to five per video, and maintaining total audiovisual durations below approximately 6 minutes, the videos operationalized evidence-based strategies for managing cognitive load. Evaluation data support this design rationale: video scripts achieved better readability scores than website text (**Supplementary Table 1**, [23]), and patients reported that videos enhanced comprehension of complex concepts such as surgical anatomy and procedural workflows [23]. However, even video scripts fell short of target readability levels (**Supplementary Table 1**), highlighting a persistent tension between clinical precision and linguistic accessibility. This suggests that future col71creation efforts should incorporate formal readability assessment as a design constraint during content development rather than an evaluation metric applied retrospectively. At the same time, readability indices remain crude proxies that focus largely on surface text features such as sentence and word length; they should therefore not be treated as definitive indicators of patient comprehensibility [34], particularly when materials are embedded in a multimedia, pathwayl71aligned intervention and have been tested directly with patients. Recent studies also suggest that generative artificial intelligence tools can support this process by rewriting existing patient education materials to meet recommended reading levels while largely preserving accuracy, although such outputs still require careful clinical and patient review before implementation [35, 36].

### Gaps in the co-creation process

Despite achieving its primary educational objectives, the platform’s evaluation highlighted three areas where the co-creation process revealed important limitations, pointing toward methodological expansions for future participatory design projects [23]. First, although workshop participants and professional material creators explicitly prioritized accessible language, formal readability metrics showed that most platform sections exceeded recommended 6^th^-8^th^ grade thresholds in both English and French. The evaluation cohort’s relatively high baseline health literacy may have masked this discrepancy during user testing, underscoring a critical limitation: participatory design that includes only high-literacy patient representatives risks producing materials that serve general cohorts but not more vulnerable populations. Future iterations should therefore treat readability as a non-negotiable design constraint, validated with patient panels that include individuals with documented low literacy. This represents a shift from co-creation as consensus-building among available and motivated stakeholders toward co-creation as deliberate oversampling of marginalized perspectives as a safeguard against inadvertent exclusion [17, 18, 37].

Second, the evaluation underscored limitations in digital user experience (UX) [23]. The co-creation process prioritized content architecture and narrative structure but did not systematically engage end-users in interface design and usability testing. Consequently, the platform exhibited deficits in mobile responsiveness, font sizing, subtitle clarity, and navigation intuitiveness; these issues were identified only during post-launch evaluation [23]. This reflects a common pattern in health information co-creation, where participatory methods focus on what to communicate while delegating how to deliver it digitally to technical experts. Future projects should extend participatory workflows to include iterative usability testing protocols such as think-aloud sessions, device-specific navigation testing, and accessibility evaluations with patients using assistive technologies. In this view, digital UX becomes not a technical afterthought but an explicit co-creation object requiring patient input throughout interface development [24, 38, 39].

Third, the external validation study highlighted a boundary in psychosocial outcomes [23]. Although patients reported enhanced trust in health information, greater preparedness, and perceived emotional support (particularly among those with higher baseline stress), group-level stress, anxiety, and depression scores did not change substantially after using the platform [23]. This pattern aligns with broader evidence that multimedia educational interventions grounded in cognitive learning principles are effective for improving knowledge, confidence, and perceived preparedness but may not, on their own, achieve large or sustained reductions in psychological distress [2]. In some surgical settings, structured and repeated education has been shown to reduce perioperative anxiety and depression, especially when it addresses follow-up, self-management, and expectations in a systematic way, suggesting that educational content can contribute to emotional outcomes when integrated into broader psychosocial support [9, 40, 41]. In our case, the persistence of distress despite improved knowledge underscores that informational preparedness, while necessary, is insufficient to address all emotional dimensions of surgical decision-making and recovery. More comprehensive support likely requires workflows that combine education with approaches such as systematic distress screening, nurse-led counseling, and referral to psychological services, ideally co-designed with patients, clinicians, and mental health professionals.

### Refining co-creation in patient education

Building directly on these gaps in readability, digital UX, and psychosocial scope, the Butterfly platform illustrates both the strengths and boundaries of participatory design in patient education. On one hand, the process produced credible, structured, and actionable materials that aligned with clinical workflows and patient-reported needs. On the other, it highlighted how co-creation practices could be strengthened to better address equity, accessibility, and broader patient support. First, readability should move from aspiration to explicit design requirement, with formal targets specified from the outset and revisited iteratively by panels that include patients with low literacy and other communication challenges, as recommended in participatory health literacy work. Second, co-creation should extend beyond content to the digital interface itself, systematically involving patients in assessing usability and accessibility across devices and contexts, in line with emerging guidance on co-design of digital health interventions and digital health equity [38, 39, 42, 43]. Third, the boundaries between educational and psychosocial aims should be made explicit, with complementary pathways for distress screening and referral co-designed with patients, clinicians, and mental health professionals when needs extend beyond information provision [2, 41].

Taken together, these directions reposition co-creation not as simple stakeholder consultation but as a dynamic process of managing tensions: between clinical precision and linguistic accessibility, expert knowledge and lived experience, technical feasibility and user-centered design, and informational support and psychosocial care. Making these tensions explicit and negotiable within participatory workflows is consistent with contemporary participatory design frameworks and can help future projects produce educational interventions that are evidence-based, accessible, usable, and appropriately scoped for diverse patient needs and populations [18, 19, 24].

### Study strengths and limitations

This study provides a detailed, replicable account of a multi-phase participatory process for developing patient-centered educational resources, explicitly grounded in theoretical frameworks and operationalized through structured methods. The integration of multidisciplinary clinician and patient workshops with iterative refinement and external validation strengthens both the methodological rigor and practical applicability of the approach. Unlike many thyroid education resources that provide fragmented or outdated information, the Butterfly platform delivers guideline-consistent, pathway-structured content aligned with six stages of surgical care, closely reflecting recommendations in current international guidelines for the management of thyroid nodules and differentiated thyroid cancer [21, 22]. Transparent documentation of the co-creation process, including both successes and limitations, offers a practical template for developers, clinicians, and researchers seeking to apply participatory methods in other disease areas.

The external validation cohort was relatively small and characterized by a relatively high baseline health literacy [23], which may limit generalizability to more diverse or vulnerable populations and may have reduced the likelihood of detecting readability issues during user testing. Despite explicit attention to plain language, formal readability assessments showed that both webpage text and video scripts frequently exceeded recommended grade levels, revealing a gap between design intentions and objective literacy demands. UX and digital accessibility dimensions (such as mobile responsiveness, navigation, and subtitles) were not systematically co-designed or usability-tested, constraining conclusions about real-world digital accessibility across devices and contexts. The intervention was conceived and evaluated as an educational platform rather than as a psychosocial program; although informational outcomes were positive, stress, anxiety, and depression scores did not substantially improve at the group level [23], indicating that education alone is insufficient to address the emotional dimensions of surgical care. Finally, although the co-creation process included a patient representative from a national association, broader patient diversity (including, for example, individuals with low literacy, limited digital access, or non-dominant language backgrounds) was not systematically incorporated into design workshops, which may reduce the platform’s resonance among some underserved groups.

## CONCLUSION

Col71creating the Butterfly multimedia patient education platform with clinicians, industry partners, and a patient representative showed that participatory design can yield pathwayl71aligned, guidelinel71consistent digital education materials for thyroid surgery. The resulting webpages, animated videos, and leaflets supported patients’ informational needs and perceived preparedness but also revealed persistent challenges around readability, digital usability, and psychosocial impact that education alone cannot fully resolve. Future participatory digital patient education projects should treat readability as a nonl71negotiable design constraint, extend col71creation to systematic usability testing of digital interfaces, and explicitly define whether the intervention targets informational, psychosocial, or combined outcomes so that complementary support pathways can be col71designed where needed.

## SUPPLEMENTARY MATERIAL

**Supplementary Table 1.** Readability analysis results of the Butterfly platform’s video transcripts (English version).

**Supplementary Table 2.** Linguistic characteristics of the Butterfly platform’s video transcripts (English version) assessed using LIWC-22.

## AUTHOR CONTRIBUTIONS

MM: Contributed to the design of the study; analyzed the co-creation activities in relation to the external validation study; led the methodological framing and interpretation of results; drafted the initial version of the manuscript and prepared tables and figures.

GPS: Contributed to the conception and overall design of the study; participated in the advisory board for co-creation; supervised the external validation study and MM’s doctoral work; revised and finalized the manuscript; approved the final version and is accountable for all aspects of the work.

## Data Availability

All data produced in the present work are contained in the manuscript

## ACKNOWLEDGEMENTS

We are grateful to Marion Ribeyron from Medtronic, Inc. for her role as project lead in the cocreation of the Butterfly platform and for helpful interactions.

## FUNDING

GPS’s employing institution received a research grant from Medtronic to support the external validation study of the Butterfly platform; this grant partially covered MM’s PhD student salary. No specific funding was received for the preparation or writing of the present manuscript.

## CONFLICT OF INTEREST

GPS served as an external advisor on a Medtronic advisory board that participated in the co-creation of the Butterfly platform; the corresponding fees were paid to his employing institution. GPS’s institution also received a research grant from Medtronic to support the external validation study of the Butterfly platform, which partially covered MM’s PhD student salary. Medtronic had the opportunity to review the manuscript for factual accuracy and compliance; the authors retain full responsibility for the content and interpretation.

## REFERENCES

1. Krist, A.H., et al., Engaging Patients in Decision-Making and Behavior Change to Promote Prevention. Stud Health Technol Inform, 2017. 240: p. 284–302.

2. Zhang, Y., et al., Digital Health Psychosocial Intervention in Adult Patients With Cancer and Their Families: Systematic Review and Meta-Analysis. JMIR Cancer, 2024. 10: p. e46116.

3. Joc, E.B., et al., Quality of life of patients with irritable bowel syndrome before and after education. Psychiatr Pol, 2015. 49(4): p. 821–33.

4. Chelf, J.H., et al., Cancer-related patient education: an overview of the last decade of evaluation and research. Oncol Nurs Forum, 2001. 28(7): p. 1139–47.

5. Bujnowska-Fedak, M.M. and P. Wegierek, The Impact of Online Health Information on Patient Health Behaviours and Making Decisions Concerning Health. Int J Environ Res Public Health, 2020. 17(3).

6. Doubleday, A.R., et al., Online Information for Treatment for Low-Risk Thyroid Cancer: Assessment of Timeliness, Content, Quality, and Readability. J Cancer Educ, 2021. 36(4): p. 850–857.

7. Patel, C.R., et al., Readability assessment of online thyroid surgery patient education materials. Head Neck, 2013. 35(10): p. 1421–5.

8. Stossel, L.M., et al., Readability of patient education materials available at the point of care. J Gen Intern Med, 2012. 27(9): p. 1165–70.

9. Altinbas, B.C. and A. Gursoy, Nurse-led web-based patient education reduces anxiety in thyroidectomy patients: A randomized controlled study. Int J Nurs Pract, 2023. 29(3): p. e13131.

10. Cade, A., et al., Differences in learning retention when teaching a manual motor skill with a visual vs written instructional aide. J Chiropr Educ, 2018. 32(2): p. 107–114.

11. Wang, S.Y., T.H. Chang, and C.Y. Han, Effectiveness of a Multimedia Patient Education Intervention on Improving Self-care Knowledge and Skills in Patients with Colorectal Cancer after Enterostomy Surgery: A Pilot Study. Adv Skin Wound Care, 2021. 34(2): p. 1–6.

12. Hansen, S., et al., The Effectiveness of Video Animations as a Tool to Improve Health Information Recall for Patients: Systematic Review. J Med Internet Res, 2024. 26: p. e58306.

13. Moe-Byrne, T., et al., The effectiveness of video animations as information tools for patients and the general public: A systematic review. Front Digit Health, 2022. 4: p. 1010779.

14. Feeley, T.H., M. Keller, and L. Kayler, Using Animated Videos to Increase Patient Knowledge: A Meta-Analytic Review. Health Educ Behav, 2023. 50(2): p. 240–249.

15. Mayer, R.E., Multimedia Learning. 2nd ed. ed. 2009, Cambridge: Cambridge University Press.

16. Waxman, J.B.G., S. J., Cognitive Theory of Multimedia Learning: Applying cognitive load theory to the design of educational multimedia. 2023, Center for Health Decision Science, Harvard T.H. Chan School of Public Health.

17. McDonald, I.R., et al., What Are the Best Practices for Co-Creating Patient-Facing Educational Materials? A Scoping Review of the Literature. Healthcare (Basel), 2023. 11(19).

18. Neuhauser, L., Integrating Participatory Design and Health Literacy to Improve Research and Interventions. Stud Health Technol Inform, 2017. 240: p. 303–329.

19. Knowles, S.E., et al., Participatory codesign of patient involvement in a Learning Health System: How can data-driven care be patient-driven care? Health Expect, 2022. 25(1): p. 103–115.

20. Arcia, A., et al., A Practical Guide to Participatory Design Sessions for the Development of Information Visualizations: Tutorial. J Particip Med, 2024. 16: p. e64508.

21. Durante, C., et al., 2023 European Thyroid Association Clinical Practice Guidelines for thyroid nodule management. Eur Thyroid J, 2023. 12(5).

22. Ringel, M.D., et al., 2025 American Thyroid Association Management Guidelines for Adult Patients with Differentiated Thyroid Cancer. Thyroid, 2025. 35(8): p. 841–985.

23. Moschofidou, M., et al., Comprehensive evaluation of Medtronic’s Butterfly platform, a new audiovisual information material for patient education and shared decision-making in surgical thyroid disease. BMC Med Inform Decis Mak, 2026.

24. Wacnik, P., S.R. Daly, and A. Verma, Participatory design: a systematic review and insights for future practice. Design Science, 2025. 11.

25. Denecke, K., et al., Key Components of Participatory Design Workshops for Digital Health Solutions: Nominal Group Technique and Feasibility Study. J Healthc Inform Res, 2025. 9(3): p. 359–379.

26. Rahimi, B., et al., A Systematic Review of the Technology Acceptance Model in Health Informatics. Appl Clin Inform, 2018. 9(3): p. 604–634.

27. Krumm, I.R., et al., Making Effective Educational Videos for Clinical Teaching. Chest, 2022. 161(3): p. 764–772.

28. Niekrenz, L. and C. Spreckelsen, How to design effective educational videos for teaching evidence-based medicine to undergraduate learners – systematic review with complementing qualitative research to develop a practicable guide. Med Educ Online, 2024. 29(1): p. 2339569.

29. Giuliani, M.E., et al., Using a Nominal Group Technique to Develop a Science Communication Curriculum for Health Professionals and Clinical Researchers. J Cancer Educ, 2023. 38(5): p. 1459–1465.

30. Brame, C.J., Effective Educational Videos: Principles and Guidelines for Maximizing Student Learning from Video Content. CBE Life Sci Educ, 2016. 15(4).

31. Gutierrez-Gonzalez, R., A. Royuela, and A. Zamarron, Student engagement in a flipped undergraduate medical classroom to measure optimal video-based lecture length. Med Educ Online, 2025. 30(1): p. 2479752.

32. Baxter, K.A., N. Sachdeva, and S. Baker, The Application of Cognitive Load Theory to the Design of Health and Behavior Change Programs: Principles and Recommendations. Health Educ Behav, 2025. 52(4): p. 469–477.

33. Dhillon, V.K., et al., Preoperative information for thyroid surgery. Gland Surg, 2017. 6(5): p. 482–487.

34. Meade, C.D. and C.F. Smith, Readability formulas: Cautions and criteria. Patient Education and Counseling, 1991. 17(2): p. 153–158.

35. Kirchner, G.J., et al., Can Artificial Intelligence Improve the Readability of Patient Education Materials? Clin Orthop Relat Res, 2023. 481(11): p. 2260–2267.

36. Gupta, M., et al., Can generative AI improve the readability of patient education materials at a radiology practice? Clin Radiol, 2024. 79(11): p. e1366–e1371.

37. Doak, L.G., C.C. Doak, and C.D. Meade, Strategies to improve cancer education materials. Oncol Nurs Forum, 1996. 23(8): p. 1305–12.

38. Malloy, J., et al., Co-design of digital health interventions with young people: A scoping review. Digit Health, 2023. 9: p. 20552076231219117.

39. Voorheis, P., et al., Maximizing the value of patient and public involvement in the digital health co-design process: A qualitative descriptive study with design leaders and patient-public partners. PLOS Digit Health, 2023. 2(10): p. e0000213.

40. Jamwal, T., et al., Does Structured Patient Education Reduce the Peri-Operative Anxiety and Depression Levels in Elective Chest Surgery Patients? A Double-Blinded Randomized Trial of 300 Patients. J Patient Exp, 2023. 10: p. 23743735231151535.

41. Sun, X., et al., Effects of a mobile health intervention based on a multitheoretical model of health behavior change on anxiety and depression, fear of cancer progression, and quality of life in patients with differentiated thyroid cancer: A randomized controlled trial. BMC Med, 2024. 22(1): p. 466.

42. Piera-Jimenez, J., et al., Cocreating Principles for Digital Health Equity: Cross-Sectional, Qualitative Study for Participatory Human-Centered Design in Catalonia. J Med Internet Res, 2026. 28: p. e84129.

43. Vial, S., S. Boudhraa, and M. Dumont, Human-Centered Design Approaches in Digital Mental Health Interventions: Exploratory Mapping Review. JMIR Ment Health, 2022. 9(6): p. e35591.

