## Supplementary material for "Co-creating the Butterfly Multimedia Patient Education Platform for Thyroid Surgery Along the Patient Pathway: Lessons for Participatory Digital Patient Education": Table 1

**Table 1.** Structure and functional mapping of Butterfly platform webpages to patient information needs and journey stages.

| Chapter | Subsection | Learning objectives | Primary information needs | Patient journey stages | Associated materials |
| --- | --- | --- | --- | --- | --- |
| 1. Understanding your condition, diagnostic and treatment options | 1.1 Understanding your condition | Explain thyroid anatomy/physiology; distinguish hypo/hyperthyroidism; define nodules (95% benign); introduce parathyroids, laryngeal nerves, lymph nodes, what is thyroid cancer and types. | Initial disease awareness, understanding thyroid function and related structures | 1 | Video 1 |
|  | 1.2 Understanding the diagnostic process | Detail diagnostic journey: endocrinologist consultation, blood tests (TSH, T4/T3, calcitonin), ultrasound, scintigraphy, FNA | Clarify multi-step workup; reduce stress from repeated tests across facilities | 1-2 | Video 2 |
|  | 1.3 Understanding your treatment options | Present management strategies: surgery types (total/partial thyroidectomy, lobectomy, isthmusectomy), alternatives (surveillance, ablation); discuss risks (bleeding, nerve injury, hypoparathyroidism) | Treatment decision-making with transparent discussion of options, risks, alternatives | 3 | Video 3 |
|  | 1.4 Questions to ask your surgeon or care team | Provide structured question checklists for patient-clinician dialogue | Address decision-making anxiety; empower active participation in consultations | 3* | — |
| 2. Getting ready for your hospitalization | 2.1 What will happen before your surgery? | Outline preoperative protocols: anesthetist meetings, blood tests, preoperative test, voice and laryngeal examination. | Clarify preparation requirements and pre-admission procedures | 4 | Video 4 |
|  | 2.2 Medication advice | Provide perioperative pharmaceutical guidance | Ensure patient safety and optimal surgical conditions through medication management | 4 | — |
|  | 2.3 Managing anxiety | Present evidence-based coping strategies: exercise, sleep routines, social support | Address pre-operative anxiety as primary psychosocial concern | 4, 6 | — |
|  | 2.4 What to pack for your hospital stay? | Organize packing by category: paperwork, medications, toiletries, clothing, devices | Reduce logistical overwhelm; support practical hospital preparation | 4 | Downloadable leaflet 1 |
| 3. Understanding your hospitalization & the surgery | 3.1 Pre-admission checklist | Operationalize tasks: fasting, medication adjustments, hygiene, transportation, home support | Address medical requirements and psychosocial dimensions of hospital entry | 4-5 | Video 4, downloadable leaflet 2 |
|  | 3.2 What will happen in hospital? | Describe day of surgery: admission, fasting, anesthesia, operation duration, recovery monitoring, discharge/in-patient stay | Provide procedural transparency for hospitalization and immediate postoperative period | 4 | Video 4 |
|  | 3.3 Understand your thyroid surgery | Thyroid surgery types (total/partial thyroidectomy, isthmectomy, neck dissection), approaches (classic, minimally invasive), lymph node dissection's role in thyroid cancer treatment, and possible complications. | Support informed consent and procedural understanding through technical transparency | 3-4 | Video 3 |
| 4. Next steps and advice for the recovery period | 4.1 What are the next steps after surgery? | Understand life after thyroidectomy, common postoperative issues: identify common issues like hoarseness (temporary, voice rest), dysphagia, hypoparathyroidism, hormone and radioactive iodine treatments, describe complication management. | Self-management guidance; set recovery expectations; support timely recognition of complications | 5 | Video 4 |
|  | 4.2 Managing pain after surgery | Provide evidence-based analgesic strategies (pain relief during, after surgery and after discharge) | Empower effective self-management of pain during early recovery and at home | 4-5 | Video 4 |
|  | 4.3 Scar care after surgery | Detail wound care, sun protection, healing timeline, warning signs | Support autonomous scar management and timely consultation for complications | 5 | Video 5 |
|  | 4.4 Managing your weight after surgery | Address metabolic changes and nutritional considerations | Support weight management and metabolic adaptation post-thyroidectomy | 5 | — |
|  | 4.5 Recommendations for the recovery period | Consolidate "do/don't do" guidance: scar care, medication adherence, exercise, diet, follow-up | Support patient autonomy in self-management throughout postoperative continuum | 5 | Videos 4 & 5, downloadable leaflet 3 |
| 5. Post-surgical treatments | 5.1 Understanding your hormone treatment | Explain lifelong replacement: levothyroxine dosing, TSH monitoring, hyper/hypothyroidism symptoms | Long-term hormone replacement therapy management and monitoring | 6 | — |
|  | 5.2 Radioactive iodine treatment | Describe RAI therapy for differentiated thyroid cancer: preparation, isolation, scans, tumor markers, surveillance | Understand specialized cancer treatment and long-term recurrence surveillance | 6 | — |

*Proposed questions also concern next stages to facilitate joint decision-making at this stage.
