## Supplementary material for "Co-creating the Butterfly Multimedia Patient Education Platform for Thyroid Surgery Along the Patient Pathway: Lessons for Participatory Digital Patient Education": Table 2

**Table 2.** Animated videos within the Butterfly platform: duration, main pathway stages addressed, and primary learning objectives.

| Video | Duration | Main patient pathway stage | Key learning objectives | Primary information need addressed |
| --- | --- | --- | --- | --- |
| 1. What is the thyroid? | 4:54 | 1: Unawareness to suspicion | Introduce thyroid anatomy, function, and nodule types; explain that most nodules (>95%) are benign | Provide reliable basic information to counter online misinformation in early disease-awareness phases |
| 2. Diagnostic process | 4:01 | 2: Diagnosis confirmation | Explain blood tests, ultrasound, scintigraphy, and FNA, including typical timelines | Reduce diagnostic uncertainty and anxiety by clarifying test procedures and scheduling across multiple appointments |
| 3. Understand the surgery: Thyroid surgery | 4:17 | 3: Decision and surgical referral | Present surgical options (total/partial thyroidectomy, lobectomy), main risks, and techniques (nerve monitoring, parathyroid preservation) | Support surgical decision-making and risk understanding through procedural transparency |
| 4. Understand your hospitalization | 6:03 | 4: Preparation, hospitalization, early recovery | Describe pre-operative preparation, hospital course, admission, general anesthesia, recovery area, immediate postoperative care, managing pain and discharge expectations | Offer practical checklists and procedural expectations to reduce pre-admission overwhelm and foster confidence |
| 5. Taking care of your scar | 2:19 | 5: Postoperative recovery | Provide practical scar care techniques, healing trajectory, sun protection guidance, and warning signs | Support self-management autonomy and adherence during recovery period |
