## Supplementary material for "Co-creating the Butterfly Multimedia Patient Education Platform for Thyroid Surgery Along the Patient Pathway: Lessons for Participatory Digital Patient Education": Table 3

**Table 3.** Downloadable leaflets and corresponding patient pathway stages.

| Leaflet title | Main content focus | Patient pathway stage | Primary purpose |
| --- | --- | --- | --- |
| What to pack for my hospital stay? | Practical packing list organized by category: paperwork (insurance details, medication lists, emergency contacts), medications with dosing instructions, toiletries, comfortable loose-fitting clothing, assistive devices; explicit guidance to leave valuables at home | 4.Preparation and hospitalization | Reduce logistical overwhelm and support practical preparation for hospitalization by through concrete, actionable guidance |
| Pre-admission checklist | Step-by-step clinical and logistical tasks: fasting protocols, medication adjustments (anticoagulants, diabetes drugs), hygiene protocols (showering), transportation planning, arrangements for post-discharge home support | 4: Preparation and hospitalization | Operationalize clinical instructions into discrete, manageable tasks; address both medical requirements and psychosocial dimensions of hospital entry |
| Recommendations for the recovery period | Structured "do/do not do" guidance: scar care with sun protection, medication timing for calcium and thyroid hormone supplements, gentle exercise to prevent stiffness, dietary recommendations, follow-up appointment attendance, considerations for radioactive iodine treatment (for select cancer patients) | 5: Postoperative recovery | Promote safe self-management, treatment adherence, and patient autonomy during early recovery and postoperative care. |
