## Supplementary Table 1 for "Co-creating the Butterfly Multimedia Patient Education Platform for Thyroid Surgery Along the Patient Pathway: Lessons for Participatory Digital Patient Education"

**Supplementary Table 1.** Readability analysis results of the Butterfly platform’s video transcripts (English version).

| **Video** | **FKGL** | **GFI** | **SMOG** | **FRE** | **U.S. Grade** | **Reading Level** |
| --- | --- | --- | --- | --- | --- | --- |
| 1. What is the thyroid? | 8.6 | 10.1 | 8.8 | 60 | 9^th^ | slightly difficult |
| 2. Diagnostic process | 11.0 | 11.9 | 10.7 | 48 | 11^th^ | fairly difficult |
| 3. Understanding the surgery: Thyroid surgery | 11.0 | 12.0 | 10.2 | 52 | 11^th^ | fairly difficult |
| 4. Understand your hospitalization | 8.8 | 10.4 | 8.7 | 64 | 9^th^ | slightly difficult |
| 5. Taking care of your scar | 9.2 | 10.0 | 8.3 | 68 | 9^th^ | slightly difficult |

The Flesch-Kincaid Grade Level (FKGL) indicates the U.S. grade level needed to understand the text, with lower scores indicating easier readability; the Gunning Fog Index (GFI) estimates the years of formal education needed to understand the text on first reading, with lower scores indicate easier readability; the Simple Measure of Gobbledygook (SMOG) estimates the years of education needed to understand a piece of writing, with lower scores indicating easier readability; the Flesch Reading Ease (FRE) formula scores text on a 100-point scale, with higher scores indicating easier readability; and Scolarius provides a comparison to other texts of the same type, with lower scores indicating easier readability.
