## Supplementary Table 2 for "Co-creating the Butterfly Multimedia Patient Education Platform for Thyroid Surgery Along the Patient Pathway: Lessons for Participatory Digital Patient Education"

**Supplementary Table 2.** Linguistic characteristics of the Butterfly platform’s video transcripts (English version) assessed using LIWC-22.

| **Video** | **words** | **analytic** | **clout** | **authentic** | **tone** | **WPS** | **BigWords** | **Dic** | **linguistic** | **function** | **pronoun** | **ppron** | **i** | **we** | **you** | **they** | **negate** |
| --- | --- | --- | --- | --- | --- | --- | --- | --- | --- | --- | --- | --- | --- | --- | --- | --- | --- |
| 1. What is the thyroid? | 781 | 63.05 | 42.32 | 30.86 | 26.97 | 14.74 | 25.35 | 82.07 | 61.72 | 50.45 | 7.81 | 3.07 | 0 | 0 | 1.79 | 1.28 | 0.77 |
| 2. Diagnostic process | 674 | 79.61 | 58.6 | 20.85 | 26.01 | 17.74 | 26.71 | 83.23 | 64.84 | 52.08 | 7.42 | 4.15 | 0 | 0 | 3.41 | 0.59 | 0.74 |
| 3. Understanding the surgery: Thyroid surgery | 747 | 80.77 | 56.03 | 16.34 | 31.25 | 20.19 | 27.71 | 87.28 | 62.78 | 52.07 | 7.23 | 3.88 | 0 | 0.13 | 3.35 | 0.13 | 0.8 |
| 4. Understand your hospitalization | 1102 | 45.48 | 98.32 | 51.53 | 38.63 | 17.49 | 20.33 | 90.65 | 72.32 | 58.08 | 12.43 | 10.07 | 0 | 0 | 9.35 | 0.64 | 0.91 |
| 5. Taking care of your scar | 427 | 58.23 | 84.58 | 51.07 | 48.81 | 20.33 | 15.93 | 84.31 | 71.19 | 52.93 | 11.71 | 7.03 | 0 | 0 | 5.85 | 0.94 | 0.47 |

All values are percentages except "words" (total word count) and "WPS" (words per sentence). The LIWC-22 summary variables include: “analytic” (analytical thinking style), “clout” (confidence and social status), “authentic” (authenticity and personal engagement), and “tone” (emotional tone from negative to positive). Linguistic dimensions include: “WPS” (measure of sentence complexity), “BigWords” (words ≥7 letters), “Dic” (dictionary words), “linguistic” (all linguistic markers), “function” (function words), “pronoun” (all pronouns), “ppron” (personal pronouns), “i” (first-person singular), “we” (first-person plural), “you” (second-person), “they” (third-person plural), and “negate” (negation words).
